# Salivary Biomarkers as Pioneering Indicators for Diagnosis and Severity Stratification of Pediatric Long COVID

**DOI:** 10.1101/2023.12.17.23300097

**Authors:** Sylwia D. Tyrkalska, Fernando Pérez-Sanz, Lorena Franco-Martínez, Camila Peres-Rubio, Asta Tvarijonaviciute, Silvia Martínez-Subiela, María Méndez-Hernández, Alba González-Aumatell, Clara Carreras-Abad, Èlia Domènech-Marçal, José J. Cerón, María L. Cayuela, Victoriano Mulero, Sergio Candel

## Abstract

Long COVID, or post-acute sequelae of SARS-CoV-2 infection (PASC), manifests as persistent and often debilitating symptoms enduring well beyond the initial COVID-19 infection. Presently, a specific diagnostic test or definitive biomarker set for confirming long COVID is lacking, relying instead on the protracted presence of symptoms post-acute infection. In this study, we examined 105 saliva samples (49 from children with long COVID and 56 controls), revealing significant alterations in salivary biomarkers. Pediatric long COVID exhibited increased oxidant biomarkers, decreased antioxidant, immune response, and stress-related biomarkers. Correlation analyses unveiled distinct patterns between biomarkers in long COVID and controls. Notably, a multivariate logistic regression pinpointed TOS, ADA2, total proteins, and AOPP as pivotal variables, culminating in a remarkably accurate predictive model distinguishing long COVID from controls. Furthermore, total proteins and ADA1 were instrumental in discerning between mild and severe long COVID symptoms. This research sheds light on the potential clinical utility of salivary biomarkers in diagnosing and categorizing the severity of pediatric long COVID. It also lays the groundwork for future investigations aimed at unraveling the prognostic value of these biomarkers in predicting the trajectory of long COVID in affected individuals.

## INTRODUCTION

The novel betacoronavirus severe acute respiratory syndrome coronavirus 2 (SARS-CoV-2) was first identified in Wuhan (China) by the end of 2019 (Zhu, Zhang et al. 2020), and it rapidly spread worldwide causing the coronavirus disease 2019 (COVID-19) global pandemic that was declared by the World Health Organization (WHO) on March 11th 2020 (Cucinotta and Vanelli 2020) and that has devastated the world for years.

COVID-19 is well known, including a huge variety of symptoms that normally appear four to five days after the moment a person first become infected, ranging from mild symptoms to critical or possibly fatal illness (Gandhi, Lynch et al. 2020, Grant, Geoghegan et al. 2020). Thus, the most common symptoms are fever, dry cough, sore throat, muscle and joint pain, fatigue, headaches, anosmia, ageusia, nasal congestion and runny nose, diarrhea, eye irritation, and breathing difficulties in moderate to severe cases (Grant, Geoghegan et al. 2020, Wiersinga, Rhodes et al. 2020). Although it seems clear that several factors, such as the health status of the patients prior to infection or the type of SARS-CoV-2 variant contracted, influence the severity of the symptoms (Dao, Hoang et al. 2021, Inui, Fujikawa et al. 2022, Tyrkalska, Martinez-Lopez et al. 2022, Yoon, Lee et al. 2023), the frequent appearance of totally different disease outcomes in apparently similar patients is still not understood at all. But it is even more difficult to understand why, while the mentioned COVID-19 acute symptoms usually disappear within the next few weeks after infection in most subjects, many acute COVID-19 convalescents experience persistent symptoms after the acute infection that has been termed as long COVID, post-COVID-19 syndrome, post-acute COVID-19 syndrome (PACS), post-COVID-19 condition, chronic COVID syndrome (CCS), or post-acute sequelae of COVID-19 (PASC) (Greenhalgh, Knight et al. 2020, Baig 2021). Even though the World Health Organization (WHO) proposed a clinical definition for post-COVID-19 through a Delphi consensus in October 2021 stating that it generally occurs three months after the onset of COVID-19, with symptoms lasting at least two months and not explained by an alternative (Soriano, Murthy et al. 2022), unfortunately, there is no consensus yet and, therefore, the nomenclature, definition, epidemiology, pathogenesis, and mechanism of long COVID are still unclear. Furthermore, as a consequence of this vagueness the prevalence of long COVID is difficult to estimate, as it depends on the syndrome definition used, the time period taken into consideration in the study, and the study population, with multiple studies showing percentages of SARS-CoV-2-infected subjects with long-term symptoms that range from less than a 15% to over a 50% (Blomberg, Mohn et al. 2021, Taquet, Dercon et al. 2021). All these gaps in knowledge of the disease, together with the difficulty in differentiating the symptoms of long COVID from those of a normal, prolonged convalescence, or of other post-infection syndromes triggered by infection and immune activation, such as post-viral fatigue (Behan and Behan 1988), makes long COVID to be a diagnosis of exclusion (Leviner 2021). In any case, the main symptoms that are usually attributed to long COVID include extreme fatigue, post-exertional malaise, cognitive dysfunction, headaches, tinnitus, palpitations, chest pains, shortness of breath, sore throat, long lasting cough, joint and muscle pain, feverishness, ageusia or dysgeusia, anosmia or parosmia, diarrhoea, sleep difficulties, skin rash, and mental health problems such as depression or changes in mood (Yelin, Wirtheim et al. 2020, Al-Aly, Xie et al. 2021, Davis, Assaf et al. 2021, Lopez-Leon, Wegman-Ostrosky et al. 2021). But all these uncertainties and long-term symptoms of COVID-19, many of them with disabling consequences, are even more dramatic when the patients who suffer from them are children, as they are in crucial stages of their development. Although the severe forms of acute COVID-19 are less common in children than in adults (Chua, Shah et al. 2021), and most of the published research on long-COVID primarily focuses on adult populations, recent meta-analyses have revealed the presence of more than 40 long COVID symptoms in children and adolescents, with a prevalence of one or more of these symptoms more than 4 weeks following a SARS-CoV-2 infection of 25.24% (Lopez-Leon, Wegman-Ostrosky et al. 2022). According to other more worrying studies, the prevalence of pediatric long COVID could even reach 66% (Fainardi, Meoli et al. 2022). In children and adolescents, the most common long-term symptoms and percentage of prevalence associated are mood symptoms (e.g., sadness, tension, anger, depression, and anxiety) (16.50%), fatigue (9.66%), sleep disorders (8.42%), headache (7.84%), respiratory symptoms (7.62%), nasal congestion (7.53%), and cognitive symptoms (e.g., less concentration, learning difficulties, confusion, and memory loss) (6.27%) (Gonzalez-Aumatell, Bovo et al. 2022, Lopez-Leon, Wegman-Ostrosky et al. 2022).

Considering all the above, it is clear that the elaboration of novel strategies and clinical guidelines for the prevention, diagnosis, follow-up, and rehabilitation of long COVID, particularly in children, is essential and for this purpose the study of possible biomarkers for this disease is of great interest. These biomarkers should ideally: (1) be measured in biological samples that are easy to collect and preferably by non-invasive methods, such as saliva; (2) be related with physiological pathways that are altered in COVID-19; and (3) be measured with technology platforms and optimized protocols for rapid analysis of the biomarkers. Saliva is an extracellular biofluid with multiple biomedical applications, with the advantage that it is easy, safe, cheap, and quick to collect, without any pain or stress (Yoshizawa, Schafer et al. 2013). Saliva has often been considered as a ‘mirror of the body’, since it is an outstanding surrogate medium that cannot reflect only the physiological and physiopathological state of the oral cavity, but also of the whole organism (Lee and Wong 2009). Although human saliva comprises 99.5% water, it presents a complex composition including many important substances, being enzymes, proteins, immunoglobulins, DNA and RNA of host origin, bacteria and bacterial products, host cells, steroid hormones, ions, and volatile compounds, the main salivary diagnostic analytes (Zhang, Xiao et al. 2009).

Among the physiological pathways that can be analyzed in saliva, there are biomarkers of redox status, immune response, and stress. Given that oxidative stress has been associated with multiple human diseases (Vona, Pallotta et al. 2021), the measurement of biomarkers that reflect the redox status of the individuals can be useful when studying the pathogenesis of a disease. Some of the most used salivary biomarkers to measure this are the total oxidant status (TOS), the advanced oxidation protein products (AOPP), the derivatives of reactive oxygen metabolites (d-ROMs), the levels of Trolox equivalent antioxidant capacity in the hydrophilic fraction (TEACH), the cupric reducing antioxidant capacity (CUPRAC), and the ferric reducing ability of plasma (FRAP) (Tvarijonaviciute, Aznar-Cayuela et al. 2018, Rubio and Cerón 2021). Regarding the evaluation of the immune response, adenosine deaminase (ADA) and its isoenzymes, ADA1 and ADA2, ferritin, and total proteins are particularly interesting. The level of ADA enzymatic activity in serum reflects the cell-mediated immunity (Baganha, Pêgo et al. 1990) because, in lymphoid tissues, ADA participates in mononuclear cell maturation from monocyte to macrophage, as well as in the differentiation of B and T lymphocytes (Sullivan, Osborne et al. 1977, MacDermott, Tritsch et al. 1980). Thus, the serum level of ADA is considered as a biomarker of situations with high levels of circulating T lymphocytes (Calis, Ates et al. 2005) and chronic inflammation (Mishra, Gupta et al. 1994), with significant increases in infection (Vijayamahantesh, Amit et al. 2016), inflammation (Santosh, Renukananda et al. 2016), malignancies (Ebrahimi-Rad, Khatami et al. 2018), and immunomediated disorders (Sari, Taysi et al. 2003). Importantly, it has already been demonstrated that total ADA (tADA) and its isoenzymes ADA1 and ADA2 can be measured accurately in saliva samples, even after the chemical inactivation of SARS-CoV-2, and that their levels are significantly increased in saliva of COVID-19 patients compared to healthy controls (Franco-Martínez, Tecles et al. 2021). Also important for monitoring the immune response is serum ferritin which, besides its role as an iron storage protein, has also been described as an essential molecule in the immune system with immunosuppressive and pro-inflammatory effects (Sharif, Vieira Borba et al. 2018). Hence, a high level of ferritin in serum is considered as a marker of inflammatory, autoimmune, malignant, or infectious status, including COVID-19 (Zandman-Goddard and Shoenfeld 2007, Vargas-Vargas and Cortés-Rojo 2020). Interestingly, ferritin levels are increased in saliva of COVID-19 patients and, even more important, those levels directly correlate with the severity of the disease (Franco-Martínez, Cerón et al. 2021). Levels of total proteins in saliva are known to be significantly increased in multiple diseases (Shaila, Pai et al. 2013, Masters, Noyce et al. 2015), which can make it a biomarker of interest. Regarding biomarkers of stress, the measurement of salivary alpha-amylase (sAA) levels can also be very informative, since it is secreted by the parotid gland in response to sympathetic-adrenal-medullary system and increases in psychological and intense physical stress situations (Nater and Rohleder 2009). Indeed, sAA levels are increased in COVID-19 patients, positively correlating with their mental stress scores (Deneva, Ianakiev et al. 2022).

The objective of this work was to study the changes in the levels of a panel of different biomarkers measured in saliva, able to provide information about redox status, immune response, and stress of children with long COVID, and the correlations between these biomarkers as well as between them and multiple demographic/clinical parameters. Based on this data, we have developed predictive models to determine whether the measurement of these salivary biomarkers would be enough to reliably discriminate between children with long COVID and healthy controls, and between children with long COVID who present a severe form of the disease and those others who only have mild symptoms. This information could be of interest for a better understanding of pediatric long COVID, and to develop novel strategies that facilitate an early treatment that reduces the sequelae of the disease.

## RESULTS

### Sample overview

A total of 105 saliva samples, 49 from children with long COVID and 56 from controls of the same age range (with average ages of 13.3 and 11.6 years, respectively), were collected and included in this study for analysis. Demographic and clinical information of patients with long COVID was gathered as well, aiming to check any potential associations between these data and the different salivary biomarker levels measured in this work. Thus, data about previous relevant diseases obtained from the patient personal health records, as well as clinical condition measured after the diagnosis of long COVID, were collected to monitor the development of the disease (Table 1). Consistent with previous studies suggesting that pediatric long COVID may have an important impact on health (Lopez-Leon, Wegman-Ostrosky et al. 2022), our analysis of clinical data from children with the disease enrolled in this study revealed the great threat that long COVID poses to their health and normal development. Indeed, as many as 28.6% of them presented a severe form of the disease, including highly disabling symptoms such as extreme fatigue (44.9%), CNS symptoms (79.6%), brain fog (59.2%), respiratory symptoms (59.2%), cardiovascular symptoms (65.3%), and insomnia (46.9%) (Table 1).

**Table 1.** Long COVID patient demographics and clinical information.

|  |  |
| --- | --- |
| Number of patients | 49 |
| Average diagnosis age (range) | 13.3 (9-18) |
| Female sex (%) | 34 (69.4) |
| Mild long COVID (%) | 35 (71.4) |
| Severe long COVID (%) | 14 (28.6) |
| Relatives with long COVID (%) | 15 (30.6) |
| Previous psychiatric disorder (%) | 5 (10.2) |
| Atopy (%) | 7 (14.3) |
| Asthma (%) | 4 (8.2) |
| Fatigue test with 'high' or 'very high' result (%) | 22 (44.9) |
| CNS symptoms (%) | 39 (79.6) |
| Brain fog (%) | 29 (59.2) |
| Weight loss (%) | 11 (22.4) |
| Respiratory symptoms (%) | 29 (59.2) |
| Cardiovascular symptoms (%) | 32 (65.3) |
| Insomnia (%) | 23 (46.9) |
| Altered school attendance (%) | 14 (28.6) |
| Reduced school performance (%) | 22 (44.9) |
| Interruption of extracurricular activities (%) | 24 (49.0) |
| Interruption of sports activity (%) | 26 (53.1) |

### Salivary biomarker levels are altered in children with long COVID

The levels of 13 biomarkers were measured in saliva of children with long COVID and controls. These biomarkers were selected based on the potentially high relevance of the information that they were presumably able to provide us about the redox status (TOS, AOPP, D-ROMS, TEACH, CUPRAC, and FRAP), the immune response (ADA1, ADA2, tADA, ferritin, and Ig-RBD), the acute stress levels (sAA), and the general homeostatic status (total proteins) of children included in this study.

As a first and simple approach to check whether salivary biomarker levels were altered in children with long COVID compared to controls, these levels were analyzed jointly. For this, we performed Principal Component Analysis (PCA) to visualize the biomarker level patterns of all samples in a low-dimension space based on the salivary level matrix of the 13 biomarkers across the 105 samples. Thus, the top two principal components explained 27.1% and 23.7% of the variance of the original data, respectively (Figure 1A). As shown in Figure 1A, the samples apparently did not form two clearly different clusters as they appeared mostly intermixed, both centroids were not far apart from each other, and the confidence ellipses chiefly overlapped. However, a closer look to these PCA results revealed that while most samples located in the upper left quadrant corresponded to children with long COVID and just a few of them to controls (18 vs. 6, respectively), exactly the opposite situation was observed in the lower right quadrant (6 vs. 24, respectively) (Figure 1A). Therefore, although samples from long COVID patients and those from controls did not form two clearly separate clusters, PCA results suggested that there could be significant differences in salivary biomarker levels between both groups that deserved further research.

**Figure 1.**
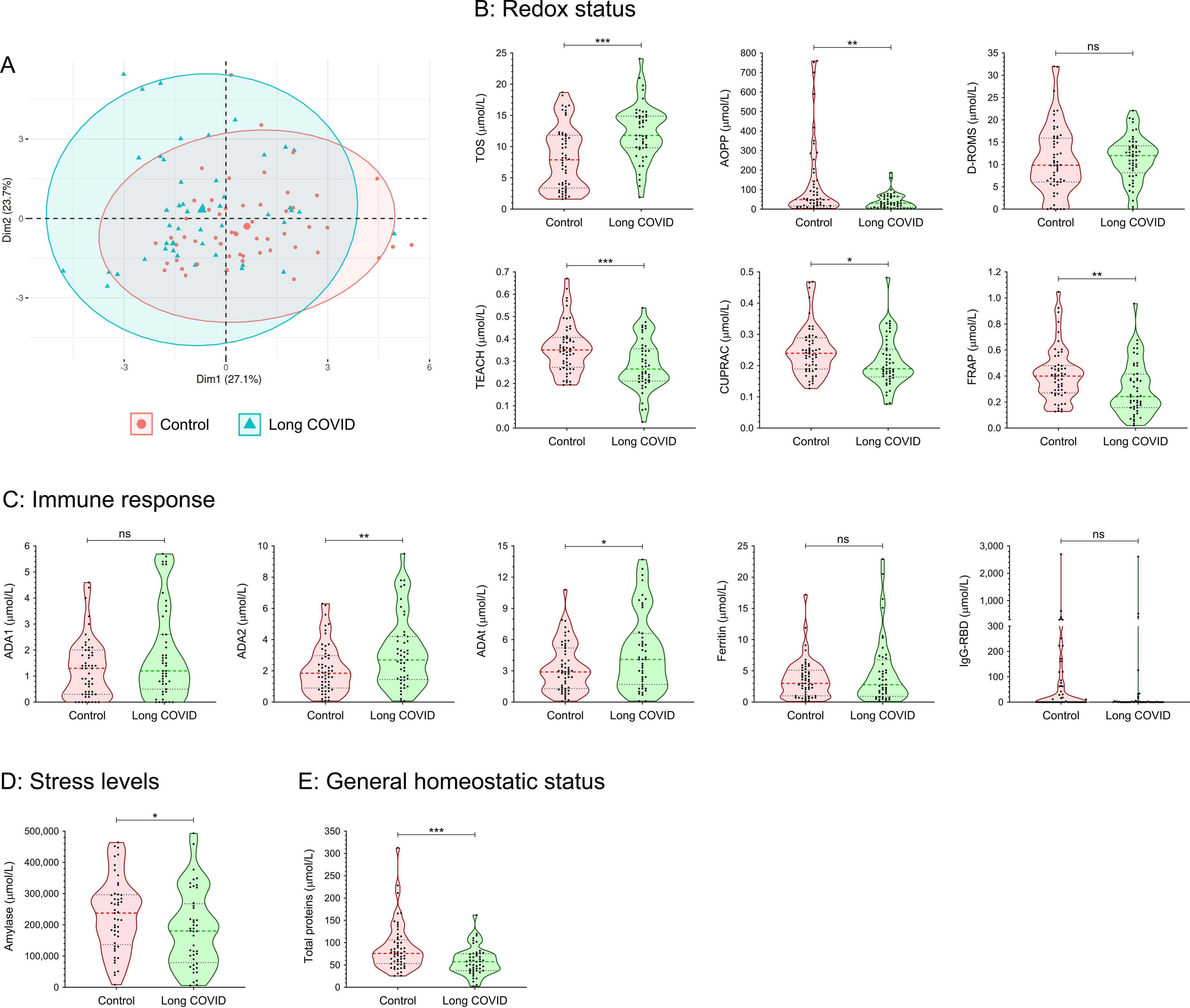
Sample overview using PCA and salivary biomarker levels. **A** Using the levels of the 13 salivary biomarkers included in this study measured in the 105 saliva samples as input, the data were linearly transformed and visualized in two-dimensional space. Each sample is represented by one dot, and colored and shaped according to the group to which it belongs. The percentage of the variance of the original data explained by each of the two principal components is indicated in the axis labels. The 90% confidence data ellipses are shown for each group. **B-E** Violin plots showing the levels of the 13 biomarkers included in this study in saliva of children with long COVID and controls. Salivary biomarkers have been grouped according to the information they provide as indicated. Each sample is represented by one dot. *P* values were calculated using Student’s t-test. ns, not significant; *P ≤ 0.05, **P ≤ 0.01, and ***P ≤ 0.001.

Our moderately promising preliminary foray into the study of the salivary biomarker levels of children with long COVID and controls by PCA (Figure 1A), led us to go further and analyze in detail the differences between both groups of children for each of the 13 salivary biomarkers included in this study individually. Interestingly, we found that while the levels of the oxidant biomarkers TOS and D-ROMS were increased in children with long COVID compared to controls, exactly the opposite result was observed for the antioxidant biomarkers TEACH, CUPRAC, and FRAP, whose levels were significantly reduced in children with long COVID (Figure 1B). Curiously, the only salivary biomarker of this group that did not follow the same pattern was AOPP, as its level was increased in children with long COVID albeit it is a measurement of the oxidative protein products (Figure 1B). Furthermore, it was surprising to observe that the levels of only two of the five biomarkers measured to evaluate the immune response, ADA2 and tADA, were significantly altered in saliva of children with long COVID compared to controls (Figure 1C). Finally, the sAA and total protein levels, that usually increased in disease (Nater and Rohleder 2009, Shaila, Pai et al. 2013, Masters, Noyce et al. 2015, Deneva, Ianakiev et al. 2022), were significantly reduced in children with long COVID compared to controls (Figures 1D and 1E).

### Correlation patterns between the different salivary biomarkers are altered in children with long COVID

After finding clear differences when comparing the salivary biomarker levels of children with long COVID and controls in many cases (Figures 1B-1E), and aiming to obtain more information which could help us understand better the biological relevance of such differences, we checked whether there were significant correlations between the levels of the different salivary biomarkers included in this study. For this purpose, we used the Pearson correlation coefficient to measure the lineal correlation for all the possible biomarker pairs, and the magnitudes (colors) and statistical significances (asterisks) of all these correlation analyses were displayed in a diagram to facilitate the joint interpretation of the results (Figure 2A). As expected, the highest correlations were those found between salivary biomarkers that are functionally related, such as the high positive correlations observed for the pairs FRAP-TEACH, CUPRAC-TEACH, FRAP-CUPRAC, ADA1-tADA, and ADA2-tADA (Figure 2A). Curiously, while some biomarkers presented significant correlations with many others, as in the cases of TEACH and the total amount of proteins in saliva, whose levels significantly correlated with 7 of the other 12 biomarkers in both cases, others significantly correlated with only a couple of the other biomarkers (TOS) or even with none of them (IgRBD) (Figure 2A).

**Figure 2.**
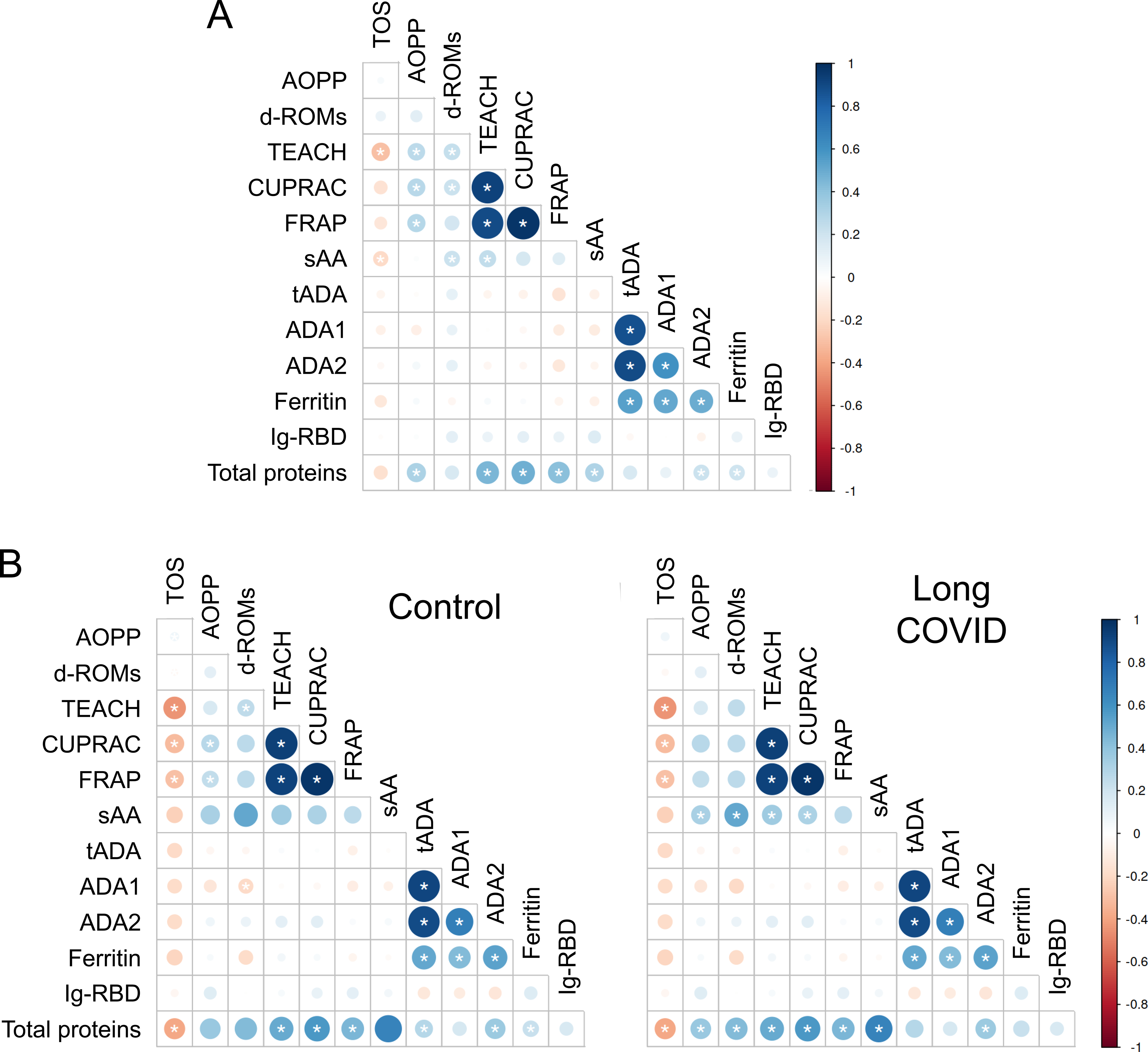
Correlations between the salivary biomarkers measured in this work. Diagrams showing the correlations between all the possible salivary biomarker pairs according to the Pearson correlation coefficients, using the salivary biomarker levels of all the children included in this study (**A**), only of control children (**B**), or only of children with long COVID (**C**). Each cell of the grid refers to the correlation between the two biomarkers faced. The circle sizes are proportional to the strength of the correlations, while the circle colors show the values of the Pearson correlation coefficients according to the legend. Asterisks denote statistically significant correlations.

The existence of significant correlations between salivary biomarkers in several cases (Figure 2A), together with the fact that one of our main goals is to be able to discriminate between children with long COVID and controls reliably, led us to wonder if both groups of children could present different correlation patterns. Thus, we decided to repeat the same analysis of correlations but this time for controls and children with long COVID separately, and results were represented in two diagrams to be able to visually compare them more easily (Figure 2B). These analyses revealed that the total number of salivary biomarker pairs with a significant correlation was almost equal in both groups of children (24 vs. 23, respectively) (Figure 2B). However, a closer look at the results showed that several correlations observed in control children, particularly those between the salivary biomarkers measured to evaluate the oxidative status (i.e. TEACH-D-ROMS, CUPRAC-AOPP, or FRAP-AOPP), were no longer statistically significant in children with long COVID (Figure 2B). In the opposite direction, the case of sAA drew special attention as it went from not significantly correlating with any other salivary biomarker in controls to doing so with AOPP, D-ROMS, TEACH, and CUPRAC in children with long COVID (Figure 2B).

In summary, these results demonstrate that not only the levels of most of the salivary biomarkers included in this study are altered in children with long COVID compared to controls (Figures 1B-1E), but also that the correlation patterns between such biomarkers are significantly different when comparing both groups of children (Figure 2B). Therefore, both types of differences could potentially be useful to discriminate between children with long COVID and those who do not suffer from the disease.

### Salivary biomarker levels are useful to reliably discriminate between children with long-COVID and controls, and also between children with mild and severe long COVID symptoms

Given that levels of 9 of the 13 salivary biomarkers measured in this study were altered in children with long COVID (Figures 1B-1E), and that the correlation patterns between the different biomarkers were also significantly distinct in these children in many cases (Figure 2B), we decided to study in detail how useful these salivary biomarkers could really be in discriminating between children with long COVID and controls. Hence, a multivariate logistic regression analysis was performed to determine which of our 13 salivary biomarkers were the most explanatory variables regarding the differences between both groups of children. Importantly, this analysis showed that TOS, ADA2, total proteins, and AOPP, in combination, were the variables that best discriminated between children with long COVID and controls. Indeed, consistent with the idea that these four salivary biomarkers could be useful to discriminate between both groups of children and contrary to the observed in the previous PCA that included all the biomarkers (Figure 1A), a new PCA analysis performed with a matrix containing the levels of only these 4 salivary biomarkers across our 105 samples resulted in two clearly different distribution patterns when comparing children with long COVID and controls (Figure 3A). In this case, the top two principal components explained 36% and 25.8% of the variance of the original data, respectively, and the confidence ellipses were distinct (Figure 3A). To further confirm the utility of TOS, ADA2, total proteins, and AOPP, combined, to discriminate between children with long COVID and controls, we elaborated a predictive model with these four salivary biomarkers and tested it. Thus, 85% of all our samples were used to elaborate and train the predictive model, while the remaining 15% of samples were utilized to test it, finding that the model presented very high accuracy, 86.67% (95% CI: 0.5954, 0.9834), with a sensitivity of 0.875 and a specificity of 0.857. Furthermore, this predictive model was also evaluated by performing a receiver operating characteristic (ROC) curve, which is a graphical plot that illustrates the diagnostic ability of a binary classifier system as its discrimination threshold is varied (Fawcett 2006), resulting in an outstanding area under the curve of 0.906 and, therefore, confirming the strength and potential utility of our predictive model (Figure 3B).

**Figure 3.**
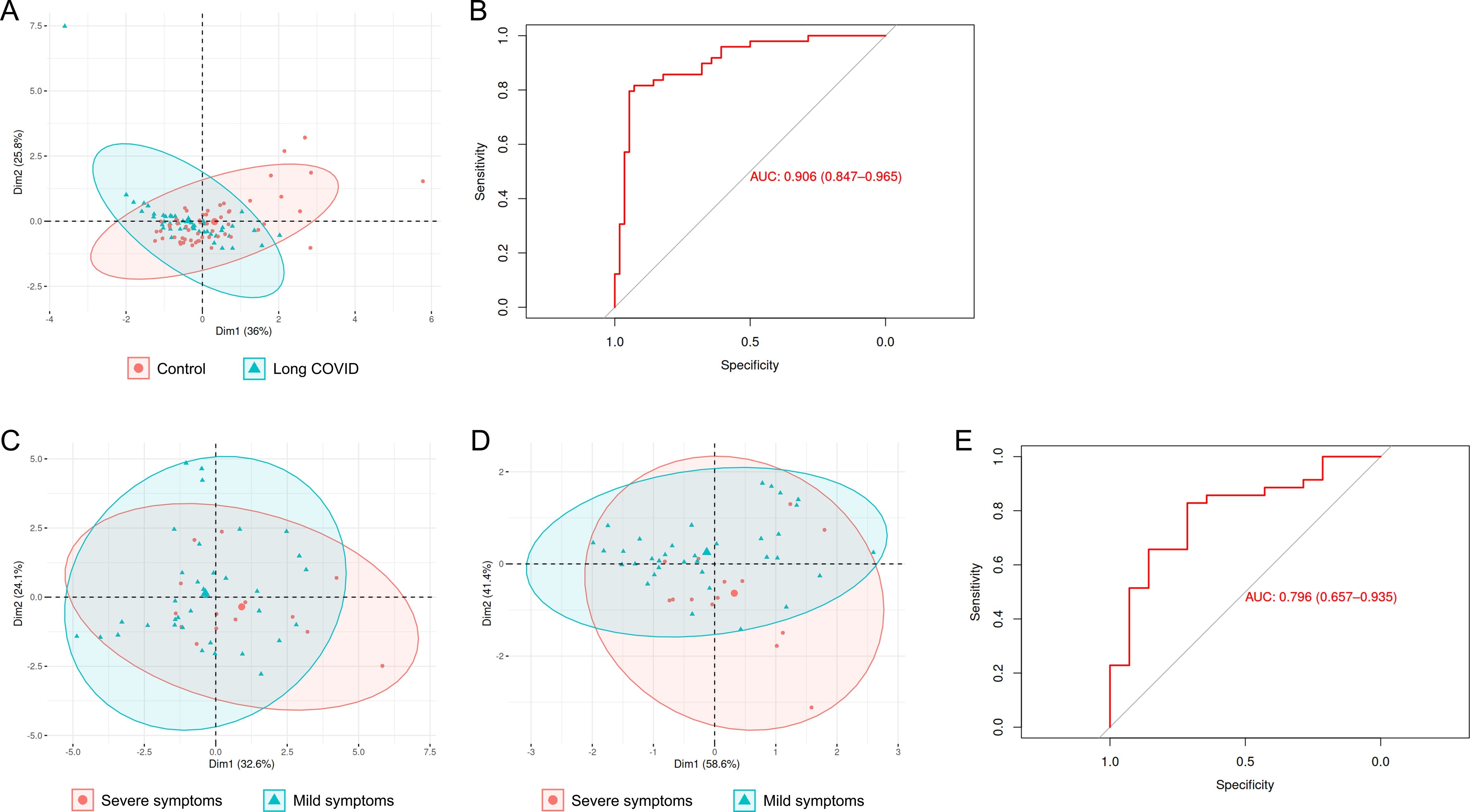
Checking the utility of the salivary biomarkers to discriminate between children with long COVID and controls, as well as between children with long COVID who present a severe form of the disease and those others with mild symptoms. **A, C, D** PCAs using as input the levels of the biomarkers TOS, ADA2, total proteins, and AOPP measured in the 105 saliva samples included in this study (**A**), the levels of the 13 biomarkers included in this study measured in the 49 saliva samples from children with long COVID (**C**), and the levels of the biomarkers total proteins and ADA1 measured in the 49 saliva samples from children with long COVID (**D**). In all three cases, the data were linearly transformed and visualized in two-dimensional space. Each sample is represented by one dot, and colored and shaped according to the group to which it belongs. The percentage of the variance of the original data explained by each of the two principal components is indicated in the axis labels. The 90% confidence data ellipses are shown for each group. **B, E** ROC curves to assess the predictive value of the levels of the salivary biomarkers TOS, ADA2, total proteins, and AOPP to discriminate between children with long COVID and controls (**B**), and of the salivary biomarkers total proteins and ADA1 to discriminate between children with long COVID who present a severe form of the disease and those others with mild symptoms (**E**).

The exceptional finding that 4 of the salivary biomarkers measured in this study could be used to reliably discriminate between children with long COVID and controls, prompted us to check, in a similar way, whether we were able to focus on the group of children with long COVID and use the biomarkers to discriminate between children who present a severe form of the disease and those others who only have mild symptoms. In this case, the multivariate logistic regression analysis using the forward and backward stepwise regression method (Efroymson and MA 1960) revealed that total proteins and ADA1, in combination, were the variables that best discriminated between children with mild and severe long COVID symptoms. Accordingly, whereas a PCA analysis performed with a matrix containing the levels of the 13 salivary biomarkers measured in this study across the 49 samples from children with long COVID resulted in two mostly overlapping confidence ellipses (Figure 3C), another PCA analysis performed with a matrix containing only the levels of total proteins and ADA1 across the same 49 samples showed two more different confidence ellipses as expected (Figure 3D). To further confirm the utility of total proteins and ADA1, combined, to discriminate between children with mild and severe long COVID symptoms, we elaborated a predictive model with these two salivary biomarkers and tested it as previously explained. This time, the model presented an accuracy of 71.43% (95% CI: 0.29, 0.96), with a sensitivity of 0.50 and a specificity of 0.80. Finally, the ROC curve for this predictive model showed an area under the curve of 0.796, suggesting that our model has good discriminatory ability (Figure 3E).

All these data together strongly suggest that the combination of the salivary biomarkers TOS, ADA2, total proteins, and AOPP could be useful to reliably discriminate between children with long COVID and controls, while the combination of total proteins and ADA1 could provide us with a tool to discriminate between children with mild and severe long COVID symptoms. Crucially, both findings could have a huge potential usefulness at the diagnosis and prognosis levels.

### Salivary biomarker levels correlate with different demographic and clinical parameters in children with long COVID

After finding that levels of the salivary biomarkers could be useful to discriminate between children with long COVID and controls, and between children with mild and severe long COVID symptoms, we aimed to investigate the potential correlations among the different salivary biomarker levels and the demographic and clinical parameters available from children with long COVID included in this study. Results of the ability of each biomarker to predict the different demographic and clinical parameters were represented in a diagram to be able to visualize them more easily (Figure 4). Interestingly, while several parameters such as the fatigue test results correlated with the levels of multiple salivary biomarkers (8), others such as the gender only correlated with the levels of one salivary biomarker (TOS) (Figure 4). From the point of view of biomarkers, the case of total proteins drew attention, since its level correlated with 8 demographic and clinical parameters, 5 of them in a statistically significant manner (Figure 4).

**Figure 4.**
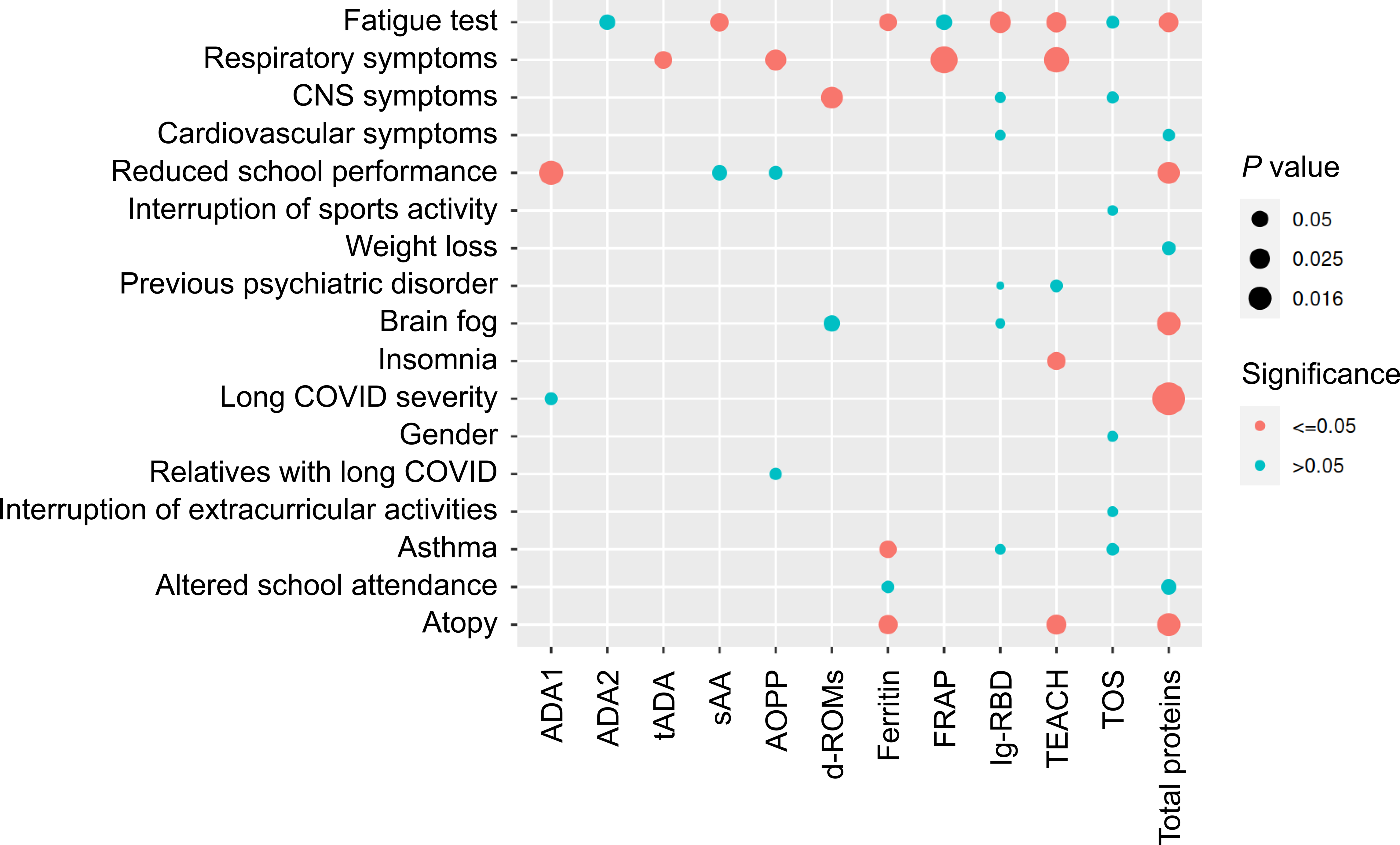
Correlations between the salivary biomarkers and the demographic/clinical variables in children with long COVID. Diagram showing the correlations between the salivary biomarkers and the demographic/clinical variables in children with long COVID, according to the Pearson correlation coefficients. Each intersection of the grid refers to the correlation between the two variables faced. The circle sizes show the strength of the correlations according to their *P* values, while the circle colors refer to the statistical significance of the correlations considering a *P* value ≤ 0.05 as the threshold.

## DISCUSSION

In this study, we have tried to shed light on the still lacunar knowledge on pediatric long COVID by taking advantage of our invaluable saliva samples from children diagnosed with the disease, together with an equally precious set of demographic and clinical data associated with such samples. The great relevance of pediatric long COVID and, therefore, the need to study it, comes not only from its high incidence as it appears in more than 25% of children infected by SARS-CoV-2 (Lopez-Leon, Wegman-Ostrosky et al. 2022), but also from the fact that more than 28% of children with long COVID present serious symptoms as consequence of this disease, according to our data (Table 1). Even though the most worrying consequence of long COVID in children is the prevalence of symptoms affecting so important systems as the central nervous (79.6%), the respiratory (59.2%), and the cardiovascular (65.3%) systems, among others, it is also alarming that a high percentage of these children see their school attendance (28.6%), school performance (44.9%), and participation in extracurricular activities (49.0%) altered (Table 1). Therefore, all this together could cause serious and difficult to recover developmental alterations in children.

The measurement of the levels of several salivary biomarkers to check whether children with long COVID were suffering from oxidative stress, revealed significant changes but reflect an apparent state of balance. Thus, while the levels of the antioxidant biomarkers TEACH, CUPRAC, and FRAP decreased in saliva of children with long COVID compared to controls, the opposite trend was observed for the oxidant biomarkers TOS and d-ROMs. The only exception was the oxidant biomarker AOPP whose levels decreased in children with long COVID, probably because the total protein levels also decreased in saliva of these children compared to controls. Salivary tADA, ADA1, and ADA2 levels provided relevant information regarding the immune response that was underway in children with long COVID. Given the prominent role of ADA in mononuclear cell maturation from monocyte to macrophage and in the differentiation of B and T lymphocytes (Sullivan, Osborne et al. 1977, MacDermott, Tritsch et al. 1980, Baganha, Pêgo et al. 1990), the increased levels of tADA and ADA2 in saliva of children with long COVID suggested that cell-mediated immunity could be more important than humoral immunity in the response of children against long COVID. Intriguingly, ADA1 levels did not change in saliva of children with long COVID. However, this is not so strange since it has recently been reported that tADA and ADA2 levels change in serum of individuals 8 weeks after diagnosis of acute COVID-

19 by PCR, whereas ADA1 levels do not change (Jedrzejewska, Kawecka et al. 2023). Furthermore, it is also not surprising that in this mentioned study the levels of tADA and ADA2 in serum decreased (Jedrzejewska, Kawecka et al. 2023) while in our work they increased in saliva, as it has been previously shown that tADA and its isoforms can respond to inflammation differently in serum or saliva (Contreras-Aguilar, Tvarijonaviciute et al. 2020). Although it has previously been reported that ferritin levels are dramatically increased in saliva of patients with acute COVID-19 (Franco-Martínez, Cerón et al. 2021), we did not find salivary ferritin levels to be elevated in children with long COVID compared to controls. Therefore, the potentiality of salivary ferritin levels to differentiate between the acute and the chronic phases of COVID-19 in children deserve further research. Surprisingly, we found that sAA and total protein levels were reduced in saliva of children with long COVID compared to controls, while both biomarkers use to be increased in disease (Nater and Rohleder 2009, Shaila, Pai et al. 2013, Masters, Noyce et al. 2015, Deneva, Ianakiev et al. 2022). Nevertheless, our sAA results are consistent with previous studies showing that sAA levels decrease in both, rats and children, with chronic stress (Wolf, Nicholls et al. 2008, Matsuura, Takimura et al. 2012). The reduced levels of total proteins observed in saliva of children with long COVID are more difficult to explain, and further research on this would require delving deeper into the eating habits of these children, since it could simply be due to a reduction in their intake.

Given that one of the main objectives of this work was the development of some simple, fast and cheap methods capable of discriminating between children with long COVID and controls, using samples that can also be obtained quickly, cheaply and minimally invasive, we did not limit our study to the measurement of salivary biomarkers individually, which could also present a high intrinsic variability between different subjects. Therefore, in addition to study changes in the levels of the different biomarkers separately, we also analyzed the correlations between all biomarker pairs to identify any correlation patterns that could exist and be considered as a characteristic signature of children with long COVID. If this signature exists, it could allow us to differentiate these children from controls through a simple correlation analysis, and probably without the need to measure more than a few biomarkers. The analysis of correlations including all samples, both from children with long covid and controls, helped us to verify that, as expected, the highest correlations were those between functionally related biomarkers (CUPRAC-TEACH, FRAP-TEACH, FRAP-CUPRAC, ADA1-tADA, ADA2-tADA, among others). Although expected, this result was still important because it served as a control to show us that our correlation analyses were reliable. Interestingly, the comparison between the salivary biomarker correlation patterns of children with long COVID and controls revealed important differences, highlighting the fact that sAA levels went from not correlating with any other biomarkers in control children to significantly correlating with the levels of other 5 biomarkers (AOPP, DROMS, TEACH, CUPRAC, and total proteins) in children with long COVID. Interestingly, when we incorporated to our correlation analyses a group of demographic/clinical variables from the children with long COVID included in this study, we found significant correlations between the levels of the salivary biomarker and these variables in 19 cases. Particularly remarkable were the cases of ferritin, TEACH, and total proteins because they were the biomarkers whose levels correlated with more of the demographic/clinical variables (with 3, 4, and 5, respectively). Importantly, we found a significant correlation between the salivary ferritin levels and the fatigue test, supporting our previously mentioned idea that iron levels might be altered in children with long COVID. Moreover, these analyses also revealed significant correlations between the sAA levels and two of the demographic/clinical variables: the school performance and the fatigue test. This is consistent with several previous studies reporting a clear association between reduced sAA levels and performance deficits during sleep loss (note that almost 47% of children with long COVID suffer from insomnia) (Pajcin, Banks et al. 2017), and between sAA levels and mental disorders in children (Jezova, Trebaticka et al. 2020).

The facts that (1) levels of most of the salivary biomarkers measured in this study (9 out of 13) were altered in children with long COVID, and (2) children with long COVID presented a characteristic pattern of correlations between the distinct biomarkers significantly different from that of control children, clearly suggested that these biomarkers could be useful for discriminating between both groups of children. These promising results prompted us to further refine our analyses by using a multivariate logistic regression analysis, finding that, among all the possible combinations of the 13 biomarkers included in this work, TOS, ADA2, total proteins, and AOPP, in combination, were the variables that best discriminated between children with long COVID and controls. But the great relevance of this discovery was not in which those 4 variables were, but in the outstanding soundness of the predictive model that included them, whose accuracy was 86.67% and with an area under the ROC curve of 0.906. Consistent with these results, our PCA analyses confirmed that the group composed only of the 4 salivary biomarkers TOS, ADA2, total proteins, and AOPP was able to discriminate between children with long COVID and controls better than using the 13 biomarkers included in this study together. This could have important implications from a practical point of view, since it meant that it would be enough to measure the levels of only those 4 biomarkers to be able to discriminate between both groups of children reliably. It would be extremely useful if the levels of the salivary biomarkers also allowed us to discriminate between children with long COVID who will present a mild form of the disease and those who will suffer severe symptoms. The multivariate logistic regression analysis performed to check this possibility showed that the group composed only of the 2 biomarkers total proteins and ADA1 was the one which best discriminated between children with mild and severe long COVID symptoms. Although these results were not as extraordinarily good as in the previous case from a statistical point of view, the predictive model including only total proteins and ADA1 had a still notable accuracy of 71.43% and an area under the ROC curve of 0.796, suggesting that they could be useful to discriminate between both groups of children. As in the previous case, the comparison of the PCA analyses including the 13 salivary biomarkers measured in this work or only total proteins and ADA1, confirmed that using only these two biomarkers we could better discriminate between children with mild and severe long COVID symptoms.

Despite the efforts of the WHO to establish a consensus that serves to clinically define long COVID (Soriano, Murthy et al. 2022), there is still no general agreement, leading to a great disparity in the criteria used to identify symptoms and diagnose the disease. As a result of these differences, not only the nomenclature, definition, epidemiology, and pathogenesis of long COVID are still unclear, but also its prevalence that varies from less than a 15% to over a 66% depending on the diagnostic criteria used (Blomberg, Mohn et al. 2021, Taquet, Dercon et al. 2021). This conundrum hinders the research into the disease pathogenesis, its approach from a clinical point of view, and the work of raising awareness among the population and institutions about its high relevance and the need to study it. Importantly, this study demonstrates that the levels of only a few biomarkers (TOS, ADA2, total proteins, and AOPP) that can be measured in saliva in a simple, fast, cheap and minimally invasive way, can be used to differentiate between children suffering from long COVID and healthy controls, or in other words, as a tool to help diagnose the disease in children following objective and standardized criteria that could be easily adopted in any hospital. In the same way, the levels of only a couple of salivary biomarkers (total proteins and ADA1) could help us to reliably and objectively classify which children with long COVID suffer from a severe form of the disease and which do not. Although this diagnostic use of the salivary biomarkers undoubtedly is very important and helpful to shed light on the field of pediatric long COVID, the ideal situation would be to be able to use them also as prognosis biomarkers to foresee which of the children infected by SARS-CoV-2 will be more likely to develop long COVID and, in addition, which of them will be more likely to develop serious symptoms of the disease. Thus, by diagnosing children at early stages of long COVID and predicting how their disease will progress, they could benefit from early monitoring and initiation of treatments and therapies relieving the severity of symptoms, thus, limiting possible alterations in their development. The results we report here are promising if we think about the possibility of using salivary biomarkers for prognosis of pediatric long COVID, albeit further research is needed to explore this possibility by measuring the levels of the distinct biomarkers in saliva of children at different times, ideally from the time of SARS-CoV-2 infection. Hence, the moment when levels of the different salivary biomarkers begin to change significantly, and the study of their kinetics is what would indicate which of the biomarkers could have a prognostic use.

This work has several strengths, many of them uncommon in this area of study: (1) it addresses a relevant but still understudied biomedical question such as pediatric long COVID; (2) collection of data was standardized in advance; (3) saliva levels of a wide variety of biomarkers were measured, providing us with information about different host responses; (4) sample sizes are very high; and (5) we have a highly valuable set of demographic/clinical data from children with long COVID. However, our study also presents several limitations that should be addressed in the future research: (1) we did not have the necessary saliva samples to measure the biomarker levels at different time points after infection of children with SARS-CoV-2, which would be useful to determine whether such biomarkers are useful for prognostic purposes and (2) we did not have a third group of children with another disease to compare and confirm that our findings are specific for long COVID. However, regarding this last issue, all our analyses were performed using the specific levels of different combinations of several biomarkers, making improbable that the same changes in the same groups of biomarkers appear in saliva of children in response to other diseases.

In summary, our study demonstrates that salivary biomarkers can be important tools to help diagnose pediatric long COVID and classify its severity in an objective and standardized manner and paves the way for future studies that explore the possibility that such biomarkers can also be used for prognostic purposes.

## METHODS

### Saliva samples and patient information collection

Saliva samples were collected from 49 children between 9 and 18 years of age, who had been diagnosed with long COVID by pediatricians at the Unidad de Covid Persistente Pediátrico of the Hospital Universitario Germans Trias i Pujol (Badalona, Spain). As a control, saliva samples were also collected from 56 healthy children within the same age range. In all cases, saliva samples were obtained under supervision using a salivette® device (Sarstedt, Germany). Participants were instructed to chew the cotton swab of the salivette® device for 1 minute and then the swab was transferred into the salivette® tube and centrifuged at 4500 rpm for 10 min. Participants were requested to refrain from eating, drinking, and performing basic oral hygiene for 1 h before sample collection. No samples showed blood contamination as determined by visual inspection. Inactivation of potential infectious viruses was performed by incubation with Np-40 to a final concentration of 0.5% for 30 minutes and were stored at −80 °C until further analysis. Regarding the inclusion criteria, participants were included in this study if they: (1) presented three or more compatible symptoms lasting longer than twelve weeks after SARS-CoV-2 infection, regardless of previous hospital admission, (2) had a positive diagnosis of SARS-CoV-2 infection or clinical suspicion (i.e., COVID-19 symptoms in participants with a close relative who had confirmation of SARS-CoV-2 infection in a situation of community transmission at the beginning of the pandemic, March 2020, when it was not possible to access to test), and (3) had persisting symptoms that were not present before COVID-19 infection. Participants were excluded if they were unable to sign the informed consent, attend the follow-up visits, or presented an alternative diagnosis. Patients who could continue with their usual activities although they presented fatigue were considered to have mild severity, those who were able to continue attending school but had to stop playing sports and extracurricular activities were considered to have moderate severity, and those who could not attend school normally were considered to have severe long COVID.

Besides the saliva samples, we collected abundant demographic and clinical information on the children with long COVID from two different sources, i.e., their clinical examinations and their patient clinical histories, aiming to facilitate the identification of any potential correlations that could exist between the biological parameters measured in the saliva samples and these demographic/clinical variables (Table 1).

### Measurement of salivary biomarker levels

Total oxidant status (TOS) was measured following the method of Erel and colleagues (Erel 2005) adapted to saliva samples. The assay is based on the oxidation of ferrous to ferric ion in the presence of various oxidant species in acidic medium. Reactive oxygen metabolites derived compounds (d-ROMs) were measured by the method described in Cesarone and colleagues (Cesarone, Belcaro et al. 1999), which is based on the capacity of transition metals to catalyse in the presence of peroxides with formation of free radicals. The trolox equivalent antioxidant capacity (TEACH) was quantified by a method based on the capacity of the the antioxidants present in the sample to reduce the ABTS, as previously reported (Barranco, Rubio et al. 2019). Cupric reducing antioxidant capacity (CUPRAC) was measured as previously described (Tvarijonaviciute, Aznar-Cayuela et al. 2017), and is based on the reduction of cupric to cuprous ion by the antioxidants present in the sample. Ferric reducing ability of plasma (FRAP) was measured in saliva following the method described by Benzie and Strain (Benzie and Strain 1996), which measures the capacity of the sample to reduce ferric to ferrous ion as previously described and validated (Tvarijonaviciute, Aznar-Cayuela et al. 2017). Salivary protein oxidation status was evaluated by measuring advanced oxidation protein product (AOPP) concentrations, as previously described by Witko-Sarsat and colleagues (Witko-Sarsat, Friedlander et al. 1996). Salivary levels of adenosine deaminase (tADA) and its isoenzymes, ADA1 and ADA2, were measured as previously described and validated (Franco-Martínez, Tecles et al. 2021). In brief, tADA was measured using a commercially available spectrophotometric assay (Adenosine Deaminase assay kit, Diazyme Laboratories, Poway, California, USA) without any treatment. Then, erythro-9-(2-hydroxy-3-nonyl) adenine (EHNA) was added to Reagent 1 and values of ADA2 and ADA1 were determined. Ferritin was measured by a commercial immunoturbidimetric assay that uses polyclonal anti-human ferritin antibodies (Tina-quant ferritin, Roche Diagnostics, Indianapolis, United States) in an automated analyzer (Olympus AU400 automated biochemical analyzer, Olympus Diagnostica GmbH, Ennis, Ireland). The assay was adapted from serum to saliva as previously reported (Franco-Martínez, Tvarijonaviciute et al. 2019). Salivary alpha-amylase (sAA) activity was measured using a colorimetric commercial kit (Alpha-Amylase, Beckman Coulter Inc., Fullerton, CA, USA) following the International Medicine (IFCC) method (1999, Rohleder and Nater 2009), as previously reported and validated (Tecles, Fuentes-Rubio et al. 2014). Total protein levels in saliva were measured using a colorimetric assay (protein in urine and CSF, Spinreact, Spain) following the manufacturer’s instructions. Salivary anti SARS-CoV-2 RBD IgG (IgG-RBD) levels were measured by a method developed and validated in the authors laboratory (Martínez-Subiela, Franco-Martínez et al. 2022). All measurements were performed in an automated analyser (Olympus AU400 automated biochemical analyzer, Olympus Diagnostica GmbH, Ennis, Ireland) except anti SARS-CoV-2 RBD IgG that were measured in EnSpireAlpha plate Reader (PerkinElmer (MA, USA).

### Bioinformatics and statistical analysis

All the bioinformatic analyses included in this study were performed with R (v.4.3.1) (https://www.R-project.org/). Principal component analyses (PCA) were conducted using the R package for multivariate analysis FactoMineR (S Lê, J Josse et al. 2008) to check, based on the levels of the different salivary biomarkers tested in each case, whether there was a significant separation between children with long COVID and control children, or between children with long COVID displaying a severe form of the disease and those others with mild symptoms. Levels of each salivary biomarker in children with long COVID and controls were represented in violin plots using GraphPad Prism (version 8.0.2 for Windows, GraphPad Software, Boston, Massachusetts USA, www.graphpad.com), and the same software was used to analyze the differences between both groups by Student’s t-test. The R package corrplot (https://github.com/taiyun/corrplot) was used to study the correlations between the levels of the different salivary biomarkers in (1) all the individuals, (2) healthy controls, and (3) individuals with long COVID, by calculating the Pearson correlation coefficient for all the possible biomarker pairs. Moreover, the same methodology was used to study the correlations between the levels of the salivary biomarkers and the demographic/clinical variables in children with long COVID. A multivariate logistic regression analysis using the forward and backward stepwise regression method (Efroymson and MA 1960) was performed to identify which variables best discriminated between children with long COVID and controls in one case, or between children with long COVID displaying a severe form of the disease and those others with mild symptoms in another case. This bi-directional stepwise procedure is a combination of forward and backward selection and elimination that tests the variables evaluated and all their possible combinations, using at every step the *t* statistics for the coefficients of the variables being considered (Efroymson and MA 1960). Once the variables that best discriminated between groups of children were identified (TOS, ADA2, total proteins, and AOPP to discriminate between children with long COVID and controls, and total proteins and ADA1 to discriminate between children with long COVID displaying a severe form of the disease and those others with mild symptoms), only those selected biomarkers were used to perform regression predictive models by using data from 85% of individuals to train the models, and from the remaining 15% of subjects to test them. These predictive models were also evaluated by performing ROC curves using the R package pROC (Robin, Turck et al. 2011). The R package ggplot2 version 3.4.3 was used to generate all the diagrams showing the results of the correlation analyses (H 2016).

## Data Availability

All data produced in the present study are available upon reasonable request to the authors.

## Acknowledgements

This work was supported by the grant 00006/COVI/20 to VM and MLC funded by Fundación Séneca-Murcia, the Saavedra Fajardo contract 21118/SF/19 to SC funded by Fundación Séneca-Murcia, the Juan de la Cierva-Incorporación contract to SDT funded by Ministerio de Ciencia y Tecnología/AEI/FEDER. The funders had no role in the study design, data collection and analysis, decision to publish, or preparation of the manuscript.

## Authors’ contributions

The authors offer the following declarations about their contributions: Conceived and designed the experiments: SC, MLC and VM. Performed the experiments: SC, SDT, FPS and SMS. Analyzed the data: SC, SDT, FPS, SMS, MMH, JJC, MLC, VM. Provided patients’ samples and clinical data: MMH, AGA, CCA, EDM. Writing-original draft: SC. Writing-review & editing: MLC, VM, MMH, AGA, SMS, JJC. All authors have read and agreed to the published version of the manuscript.

## Ethics approval and consent to participate

All procedures in this work were carried out following the principles expressed in the Declaration of Helsinki, as well as in all the other applicable international, national, and/or institutional guidelines for the use of samples and data and have been approved by the Comité de Ética de la Investigación (CEIm) at Hospital Clínico Universitario Virgen de la Arrixaca (protocol number 2020-10-12-HCUVA – Effects of aging in the susceptibility to SARS-CoV-2). In all cases, the parents or legal guardians of the children signed informed consents accepting that the saliva samples were used for research.

